# Cannabis use and psychotic-like experiences in the *All of Us* Research Program

**DOI:** 10.64898/2025.12.01.25341322

**Authors:** Emma C Johnson, Zhen Luo, Pamela N Romero Villela, Arpana Agrawal, Alexander S Hatoum, Nicole R Karcher

## Abstract

**Background and Hypothesis:** Cannabis use has been linked to psychotic-like experiences (PLEs). Amid increasing legalization, we examined the extent to which cannabis use is associated with PLEs after adjusting for other risk factors in a contemporary United States sample.

**Study Design:** We performed a cross-sectional analysis of self-reported cannabis use and four types of self-reported PLEs (auditory and visual perceptual distortions, referential ideation, and persecutory ideation) in the population-based biobank, the *All of Us* Research Program release 8 (maximum analytic N = 62,153).

**Study Results:** Cannabis ever-use (ORs = 1.21 – 1.44, p-values < 2.7e-6) and more frequent past 3-month cannabis use (within lifetime ever-users) were associated with all four PLEs (χ^2^(4) = 21.06 – 70.09, p-values = 3.08e-4 to 2.17e-14), and these associations remained when adjusting for personal and family history of schizophrenia and polygenic liability for schizophrenia. The schizophrenia polygenic score, but not cannabis use frequency, was correlated with greater likelihood of being prescribed medication for the PLEs. When adjusting for lifetime ever-use of other substances, cannabis ever-use was no longer associated with PLEs, while methamphetamine use, cigarette use, and opioid use were associated with PLEs (ORs = 1.22 to 1.65, p-values < 1.68e-05).

**Conclusions:** Prior associations between cannabis use and PLEs may have been confounded by comorbid use of other substances. Future studies that distinguish cannabis use from other substance use in the etiology of PLEs could provide insight into this transdiagnostic construct.

## INTRODUCTION

Psychotic-like experiences (PLEs) are relatively common in the general population, with one study from a representative United States sample finding that almost one third of the sample (26.7%) reported experiencing at least one PLE^1^. PLEs include auditory and visual perceptual distortions (hearing or seeing things that are not really there), referential ideation (attributing significant and personal meaning to coincidences and everyday events), and persecutory ideation (false beliefs that one is being targeted, spied on, followed, etc). While PLEs may represent trait-like variability in personality, transient variations in behavior, or divergence from cultural norms, they can also be harbingers of more serious psychopathology, being associated with a 4-fold increase in risk for psychotic disorders^2^. Risk factors for PLEs include genetic liability for PLEs and schizophrenia, adverse early life events, stress, impairments in cognition, and substance use. Cannabis use can induce acute, transient PLEs (e.g., administration of delta-9 tetrahydrocannabinol (THC), the primary psychoactive ingredient in cannabis, causes schizophrenia-like positive and negative symptoms, altered perception, and cognitive impairment^2^), and some epidemiological studies have found associations between cannabis use and general (not substance-induced) PLEs^3^. More worrisome is mounting evidence that heavy, and especially high-potency, cannabis use is linked to increased risk of psychotic disorders^4–6^, including schizophrenia. However, not everyone who uses cannabis reports PLEs or will develop a psychotic disorder. One potential pathway of risk is described by the “diathesis-stress” theory, which hypothesizes that cannabis may potentiate or accentuate genetic vulnerability, thus serving as a moderator of genetic risk.

There has been great interest in testing this diathesis-stress theory, and a previous paper by Wainberg and colleagues found that cannabis use had a dose-response effect on PLEs in a sample from the UK Biobank (N = 109,308), such that more frequent cannabis use was associated with greater likelihood of reporting PLEs^7^. They also found that cannabis use moderated the relationship between genetic liability for schizophrenia and PLEs, such that participants with greater genetic liability for schizophrenia showed a stronger association between cannabis use and PLEs. These findings are important for understanding potential risks of cannabis use and suggest that individuals with a family history of schizophrenia, for example, might be at especially high risk of experiencing adverse consequences of cannabis use. However, replication of these findings, especially cross-culturally and nationally, is needed to determine whether these results are generalizable.

Furthermore, Wainberg and colleagues were unable to investigate the effects of other substances beyond cannabis (aside from alcohol drinking frequency and smoking status), but it has been documented that use of other substances (e.g., methamphetamine) can also increase risk for PLEs^8^. Polysubstance use may further increase one’s risk for experiencing PLEs.

Finally, it may be that higher genetic liability for schizophrenia is also correlated with cannabis use (a gene-environment correlation), and this could explain previously observed interactions between polygenic scores for schizophrenia and cannabis use. It is important for studies of gene-environment interactions to examine gene-environment correlations as well.

We performed cross-sectional analyses of the relationship between cannabis use and PLEs in the *All of Us* Research Program^9^, a large biobank in the United States that has collected genomic data, survey data on substance use and PLEs, and electronic health records (EHR). Given expanding cannabis legalization in the United States, it is important to document the relationship between cannabis use and PLEs in a nation-wide sample. As PLE types can differ in terms of prevalence and, potentially, risk factors, we examined PLE types separately, as well as endorsement of any PLE regardless of type. We characterized the relationships between cannabis ever-use, past 3-month cannabis use frequency within lifetime ever-users, polygenic scores for schizophrenia, and PLEs in up to 62,010 people. Despite the difference in samples (US (N = 62,010) vs UK (N = 109,308), wider age distribution in *All of Us*, etc), we hypothesized that we would generally replicate the findings from Wainberg and colleagues in their UK Biobank analysis.

## METHODS

### Sample

The *All of Us* Research Program (AoU) is a large-scale population-based biobank based in the United States^9^. The v8 release of data includes genomic data, electronic health records (EHR), and survey responses for up to 633,540 participants. We included all participants who had survey data on cannabis use and PLEs (max analytic N = 62,010).

### Measures

#### Self-reported cannabis use and other substance use

We used data from the Lifestyle survey to create a binary variable of self-reported lifetime use of cannabis (1 = “yes”, 0 = “no”) and, within lifetime ever-users, a categorical measure of cannabis use frequency over the past 3 months, ranging from “Never” to “Daily”. We also used the Lifestyle survey to create binary variables for use of other substances, including cigarettes, alcohol, methamphetamines, stimulants, prescription stimulants, sedatives, inhalants, hallucinogens, street opioids, prescription opioids, and cocaine (**Supplemental Table S1**).

#### Self-reported psychotic-like experiences

We used data from the Behavioral Health and Personality survey to derive binary variables of endorsement of four possible lifetime PLEs: auditory perceptual distortions, visual perceptual distortions, referential ideation, and persecutory ideation, and whether a participant endorsed any of these PLE types (any PLE). We also used data from this survey on whether the PLEs were distressing, whether the participant ever talked to a medical professional about the PLEs, and whether they were ever prescribed medication to treat the PLEs. The response options for the queries regarding professional medical help or medication prescriptions were “Yes” or “No”. Distress was measured on a 5-point scale ranging from “Not distressing at all. It was a positive experience” to “Very distressing”.

#### Personal and family history of schizophrenia

Using data from the Personal and Family Health History survey, we created a binary variable representing self-reported history of a schizophrenia diagnosis in self *or* immediate family (grandparent, parents, sibling, children).

#### Polygenic liability for schizophrenia

We used PRS-CSx^10^ to generate weights based on effect sizes from two discovery genome-wide association studies (GWAS) of schizophrenia in European ancestry^11^ (N = 161,405) and African ancestry^12^ (N = 15,802) samples. We then used the --score function in PLINK to create polygenic scores (PGS) in All of Us participants of European and African genetic ancestries (as determined by the All of Us research team^13^), with these PGS representing relative genetic liability for schizophrenia.

#### Diagnoses

In the electronic health records (EHR) data, we used the International Classification of Diseases, Ninth Revision, Clinical Modification (ICD-9-CM), and International Classification of Diseases, Tenth Revision, Clinical Modification (ICD-10-CM) diagnosis codes for capturing schizophrenia diagnoses. For Bipolar affective disorder (currently, manic, severe, with psychosis) and psychotic disorder, we used SNOMED CT codes to determine the diagnosis. All ICD-9/10-CM and SNOMED CT codes were identified within the Observational Medical Outcomes Partnership (OMOP) Common Data Model framework.

#### Covariates

In primary analyses described below, we controlled for year of birth, self-identified gender, self-identified race, self-identified ethnicity, and deprivation index. In models that included the PGS for schizophrenia, we covaried for the first ten genetic ancestry principal components (specific to each ancestry).

#### Statistical analyses

Our primary analysis used a series of logistic regression models to examine whether lifetime cannabis ever-use and past 3-month cannabis use frequency within ever-users were associated with the four PLEs or any PLE (assessed as the outcome variable in independent models). Secondary analyses tested whether these associations remained after controlling for self or family history of schizophrenia or polygenic liability for schizophrenia, or after excluding participants with diagnoses of psychotic disorder, bipolar affective disorder, and schizophrenia in their EHR.

We also tested whether cannabis use frequency was associated with how distressing the PLE was reported to be, whether participants had ever talked to a medical professional about their PLE(s), and whether they were ever prescribed medication to treat their PLE(s).

We then tested whether there was an interaction effect between lifetime cannabis ever-use or past 3-month cannabis use frequency and the PGS for schizophrenia (gene-environment interactions) contributing to risk for PLEs, and whether these measures were correlated (gene-environment correlations), which can confound gene-environment interactions. In the interaction models, we also covaried for all PGS x covariate and E x covariate cross-terms, per recommendations^14^.

Finally, we also tested whether endorsement of other substances (lifetime use of cigarettes, alcohol, methamphetamines, stimulants, prescription stimulants, sedatives, inhalants, hallucinogens, street opioids, prescription opioids, and cocaine) was associated with PLEs, and whether cannabis ever-use remained associated with PLEs after adjusting for ever-use of these other substances.

All models that included the PGS were performed separately within ancestry groups, as current PGS methods generally perform poorly when ported across ancestries^15^. The ancestry-specific estimates were then meta-analyzed using a fixed-effects meta-analysis (implemented using the ‘metafor’ R package^16^) for a final cross-ancestry estimate. Across primary and secondary analyses, we performed a total of 93 tests; thus, the Bonferroni-corrected alpha level for statistical significance = 5.4e-4.

## RESULTS

Of participants who responded to the Behavioral Health and Personality survey, 6% endorsed visual perceptual distortions, 6% endorsed auditory perceptual distortions, 2% endorsed referential ideation, and 2% endorsed persecutory ideation (demographic data reported in **Table 1**). Of those who responded that they had used cannabis at least once in their life (N = 297,579) and responded to the follow-up question regarding frequency of past 3-month use (N = 143,102), 66% reported no use over the past 3 months, 16% reported using cannabis once or twice, 4% reported monthly use, 5% reported weekly use, and 9% reported daily use.

**Table 1.** Demographic characteristics of analytic sample for participants who reported using cannabis in their lifetime *and* provided data on past 3-month cannabis use and psychotic-like experiences (PLEs).

| Variable | Visual perceptual distortions | Auditory perceptual distortions | Referential ideations | Persecutory ideations | No PLEs | Total Sample |
| --- | --- | --- | --- | --- | --- | --- |
|  | (N=1413) | (N=1351) | (N=438) | (N=465) | (N=22722) | (N=25774) |
| Age (Mean (SD)) | 51.31 (16.85) | 49.70 (16.53) | 48.30 (16.51) | 46.84 (15.36) | 56.03 (16.14) | 55.72 (16.25) |
| Gender (% female) | 66.33 | 66.67 | 64.07 | 60.67 | 66.18 | 66.22 |
| Race (%) |  |  |  |  |  |  |
| White | 74.66 | 76.91 | 70.09 | 71.18 | 84.46 | 83.64 |
| Black | 11.18 | 10.36 | 15.75 | 13.98 | 5.09 | 5.65 |
| Asian | 1.2 | 1.55 | 1.6 | 2.8 | 2.23 | 2.14 |
| Middle Eastern or North African | 0.28 | 0.3 | 0.23 | 0.22 | 0.37 | 0.36 |
| Native Hawaiian or Other Pacific Islander | 0.07 | 0.15 | - | 0.22 | 0.04 | 0.04 |
| Other | 12.6 | 10.73 | 12.33 | 11.61 | 7.82 | 8.17 |
| Ethnicity (% Hispanic) | 11.38 | 9.63 | 11.27 | 11.62 | 6.07 | 6.45 |
| Deprivation index (Mean (SD)) | 0.32 (0.06) | 0.31 (0.06) | 0.32 (0.06) | 0.32 (0.06) | 0.30 (0.06) | 0.31 (0.06) |
| Cannabis ever-use (% yes) | 100 | 100 | 100 | 100 | 100 | 100 |
| Past 3-month cannabis use frequency (% yes) |  |  |  |  |  |  |
| Never | 66.67 | 67.88 | 62.56 | 62.8 | 79.3 | 78.38 |
| Once or twice | 14.58 | 14.29 | 14.84 | 13.33 | 10.4 | 10.69 |
| Monthly | 3.26 | 2.81 | 3.88 | 4.52 | 2.57 | 2.62 |
| Weekly | 4.39 | 4 | 6.16 | 6.24 | 3.43 | 3.51 |
| Daily | 11.11 | 11.03 | 12.56 | 13.12 | 4.3 | 4.8 |

Lifetime cannabis ever-use was positively associated with likelihood of all four PLEs (OR = 1.21 to 1.44, all p < 2.68e-6; analytic Ns = 61,718 - 62,153) and with endorsing any PLE (OR = 1.25, p = 1.82e-15; **Supplemental Tables S2-S6**). Frequency of cannabis use was significantly associated with all four types of PLEs (χ^2^(4) = 21.06 – 70.09, p-values = 3.08e-4 to 2.17e-14; analytic Ns = 23,769 to 23,919) and any PLE (χ^2^(4) = 76.14, p = 1.14e-15), with daily cannabis use showing the strongest association with PLEs compared to no cannabis use (**Figure 1; Supplemental Tables S7-S11**). The largest effect was seen for visual perceptual distortions, where daily cannabis users were at 2.37 greater odds of endorsing a visual hallucination compared to those who had not used cannabis during the past 3 months.

**Figure 1.**
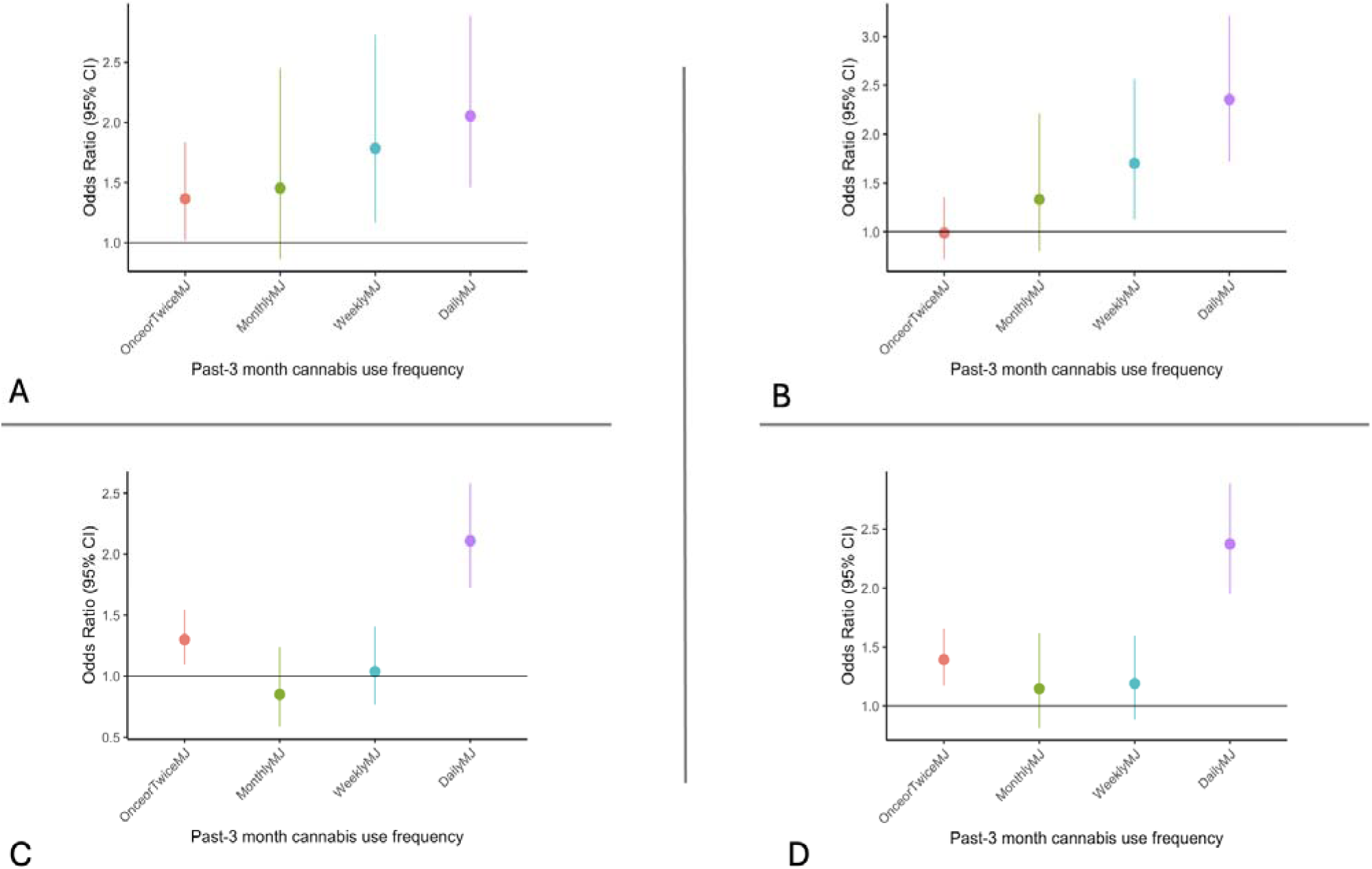
Associations between past 3-month cannabis use frequency and psychotic-like experiences. The group of participants reporting no cannabis use was the reference group for all panels. A. Associations with referential ideation. B. Associations with persecutory ideation. C. Associations with auditory perceptual distortions. D. Associations with visual perceptual distortions. OnceorTwiceMJ = using cannabis one or twice in the past three months; MonthlyMJ = using cannabis monthly in the past three months; WeeklyMJ = using cannabis weekly in the past three months; DailyMJ = using cannabis daily in the past three months.

When adjusting for family history of schizophrenia (which itself was strongly associated with likelihood of endorsing PLEs, especially persecutory ideation (OR = 6.13, p = 2.69e-18)), daily cannabis use remained associated with greater risk of all four PLEs (ORs = 2.26 to 2.9, p = 0.02 to 7.98e-5), although we note that the analytic sample sizes dropped significantly due to missing data on family history (Ns = 7,745 to 7,791). In the models that adjusted for polygenic liability for schizophrenia (meta-analyzed across ancestry groups; Ns = 15,004 to 15,111), daily cannabis use was still associated with greater likelihood of all four PLEs (OR = 2.15 to 2.62, all p < 1e-4). Within participants who endorsed experiencing a PLE, we found that frequency of cannabis use was not associated with how distressing the experience was, whether the participant sought professional help for their PLEs, or whether they were prescribed medication to treat the PLEs (χ^2^(4) = 5.21 – 7.23, p-values = 0.12 – 0.49).

To minimize the potential confounding of psychotic experiences caused by psychotic disorders, we extracted the psychotic disorder, bipolar affective disorder (current manic, severe, with psychosis), and schizophrenia codes from the EHR and replicated our primary models (analytic Ns = 22,826 to 22,968) after excluding participants with these diagnoses. Cannabis use frequency remained significantly associated with PLEs (χ^2^(4) = 20.71 – 65.72, p-values = 3.61e-4 – 1.81e-13).

No interaction effects were significant after corrections for multiple testing. The strongest interactions were observed between the schizophrenia polygenic score and lifetime cannabis ever-use, such that cannabis use had a stronger risk-increasing effect on visual perceptual distortions (p = 0.012) and referential ideation (p = 0.025) in participants with higher polygenic risk for schizophrenia. However, these interactions may have been driven by a gene-environment correlation, as cannabis ever-use was significantly correlated with the schizophrenia polygenic risk score in European ancestry participants (**Figure 2**; F_1,23_ = 42.42, p = 7.47e-11; controlling for age, gender, and principal components), such that participants who reported having used cannabis at least once in their life had higher polygenic risk scores for schizophrenia on average.

**Figure 2.**
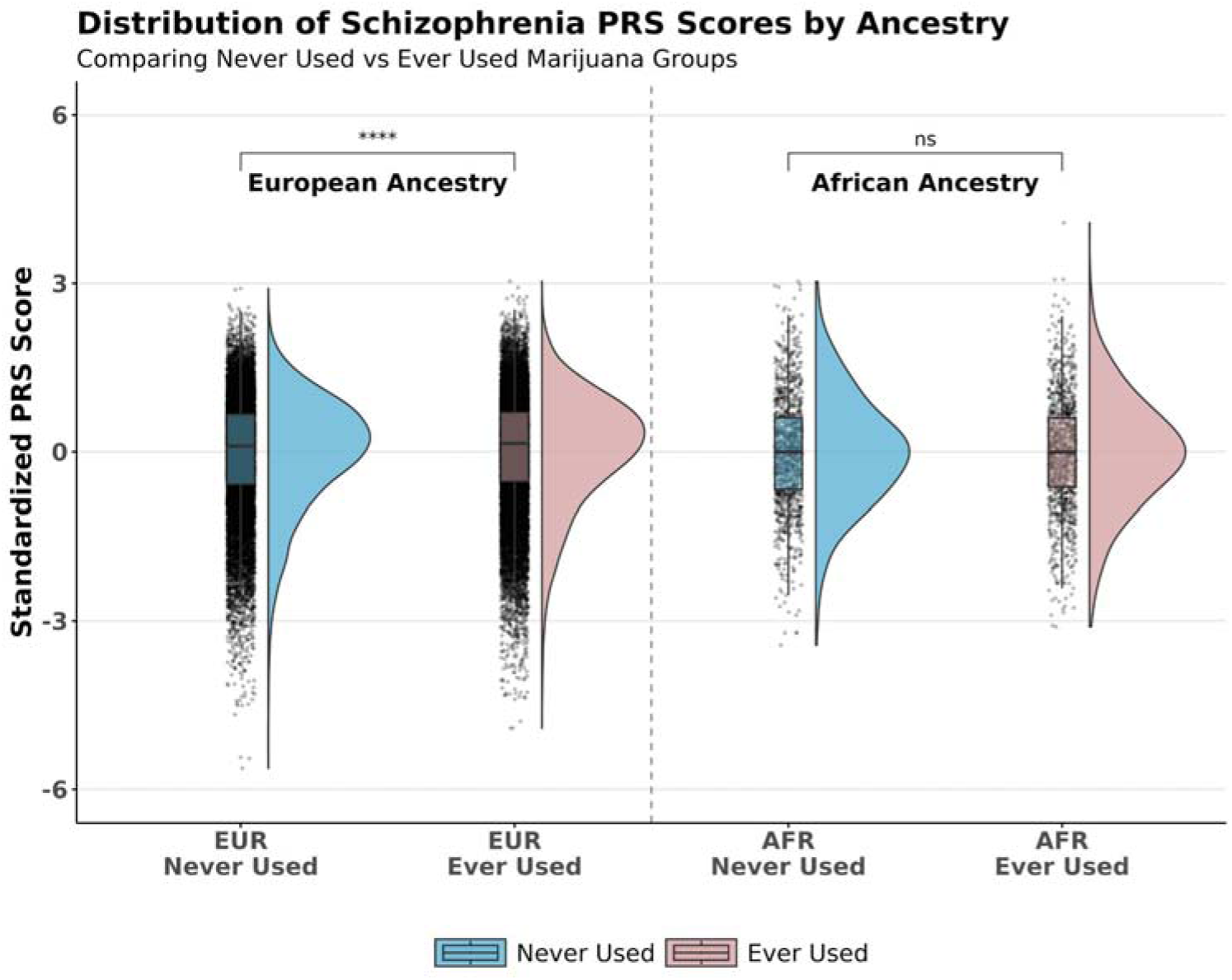
The distribution of schizophrenia polygenic liability by lifetime cannabis ever-use. p-value for association between cannabis ever-use and polygenic score in the European ancestry sample = 7.47e-11. ns = not significant (p > 0.05).

We then examined whether use of other substances was associated with PLEs, and whether adjusting for use of these other substances attenuated the relationship between cannabis use and PLEs. Each substance except for alcohol was significantly associated with risk of PLEs when each substance was tested individually, with especially strong effects for methamphetamine (OR predicting any PLE = 2.41, 95% CIs = 2.26 – 2.58; **Supplemental Tables S12-S16**). When all substance use variables were included in the same model, cannabis use was no longer associated with PLEs, but use of methamphetamines, street and street opioids, and tobacco smoking had some of the strongest associations with PLE risk (ORs = 1.32 to 1.65, p-values < 1.68e-05; **Figure 3; Supplemental Tables S17-S21**).

**Figure 3.**
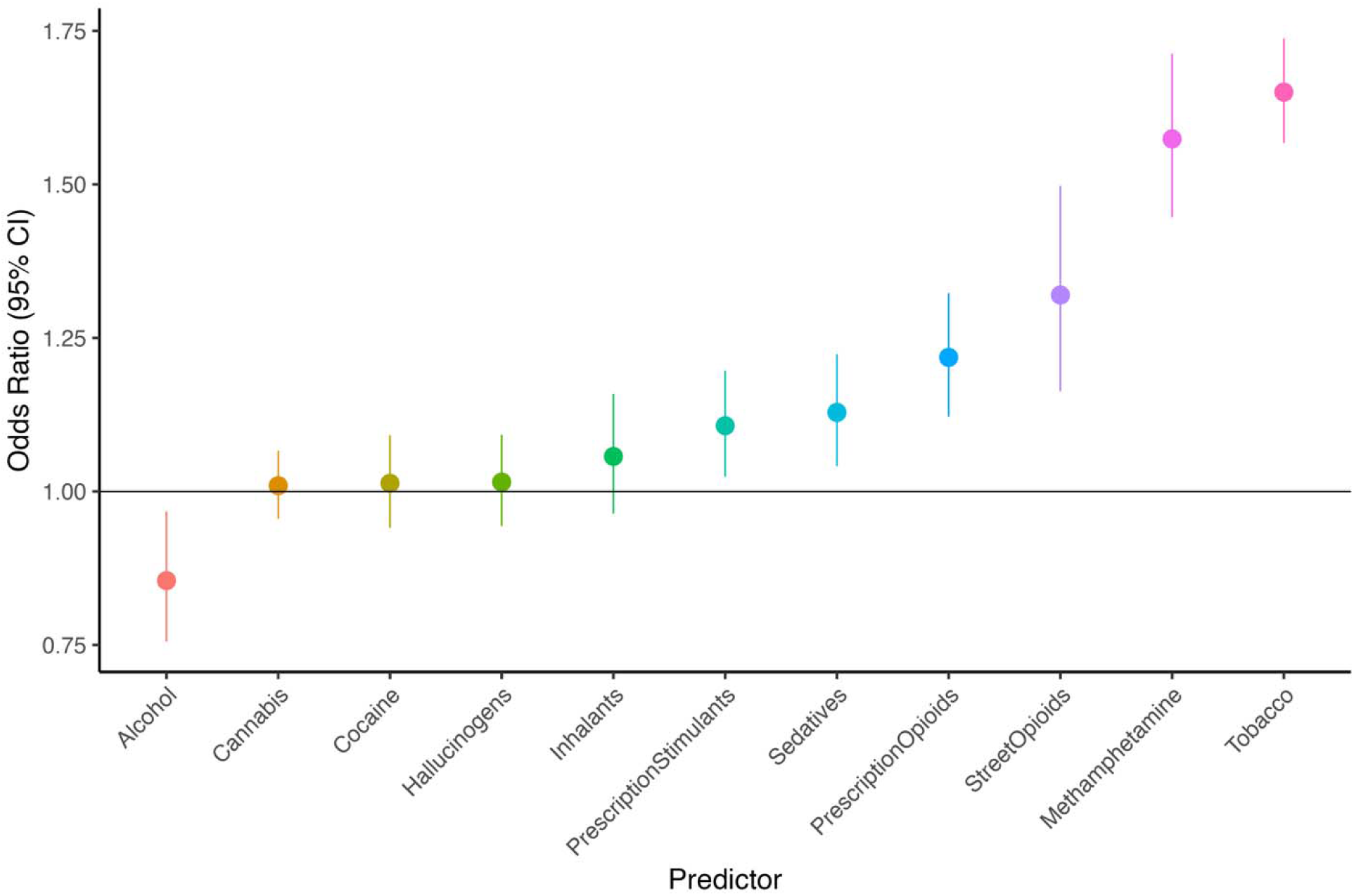
Associations between lifetime endorsement of substances and endorsement of any psychotic-like experiences, where all substance use variables were entered into the same model.

## DISCUSSION

Leveraging multi-modal data from the *All of Us* Research Program, we found robust associations between more frequent past 3-month cannabis use and likelihood of endorsing psychotic-like experiences (PLEs), including visual and auditory perceptual distortions, referential ideation, and persecutory ideation. These associations remained even when controlling for self and family history of a schizophrenia diagnosis, polygenic risk for schizophrenia, or EHR diagnoses of psychotic disorders. Two apparent (but weak) interactions between cannabis ever-use and polygenic risk for schizophrenia were likely driven by gene-environment correlations, such that participants who reported using cannabis at least once in their life tended to have higher polygenic scores for schizophrenia on average. However, when we adjusted for lifetime use of other substances, cannabis use was no longer associated with PLEs (while use of methamphetamines, opioids, tobacco smoking, and other substances were strongly associated with PLEs).

Of the four PLEs tested, we found that past 3-month cannabis use frequency had the strongest association with visual perceptual distortions, with daily cannabis users being at 2.37 greater odds of endorsing a visual hallucination compared to those who had not used cannabis during the past 3 months. Perceptual distortions, both visual and auditory, were the most commonly reported PLEs in the All of Us dataset (6%, compared to 2% for delusional ideation). This contrasts with what has generally been reported for *cannabis-induced* PLEs, where delusional ideation and paranoia are more likely to result from cannabis use than are perceptual distortions^2,17–19^. The difference in the *All of Us* PLE prevalences compared to reported prevalences of cannabis-induced PLEs in the literature may in part be due to the fact that at least one of the PLEs queried in All of Us was explicitly *not* linked to substance use – when asked about visual perceptual distortions, participants were asked to “Please do not include any times when you were dreaming or half-asleep or under the influence of alcohol or drugs.”

Interestingly, we found no evidence of an effect of past 3-month cannabis use frequency on whether participants’ PLEs were considered distressing, whether they sought professional medical help for their PLEs, or whether they were prescribed medication to treat the PLEs. This is somewhat in contrast to Wainberg and colleagues’ findings^7^, which did indicate greater distress from PLEs in cannabis ever-users compared to non-users (however, they did not assess whether frequency of use within cannabis ever-users was associated with distress). However, in a follow-up analysis, we did find that the schizophrenia PGS was associated with greater likelihood of medication being prescribed to treat their PLEs (OR = 1.26, p = 3.89e-6). This suggests that participants’ polygenic liability for schizophrenia was more tightly linked to whether the PLEs resulted in professional help-seeking and prescriptions than their cannabis intake.

When we adjusted for lifetime use of other substances, cannabis use was no longer significantly associated with PLEs. However, endorsement of use of methamphetamines, street and prescription opioids, and cigarette smoking were significantly associated with PLEs (e.g. OR for methamphetamine use predicting any PLE = 2.41, 95% CIs = 2.26 – 2.58). Wainberg and colleagues were unable to adjust for the use of other substances, and many other published examinations of risk factors for PLEs have focused solely on cannabis use or considered substance use (or use of “illicit” substances) broadly. However, methamphetamine-induced psychosis is well-documented and generally produces positive symptoms similar to those associated with schizophrenia^8^. Our findings are consistent with a wealth of literature showing that multiple substances can induce psychotic-like experiences, and it may be that some past findings of cannabis-related psychosis can be accounted for by other substances.

One limitation of this project is that our analyses were fully cross-sectional, meaning we did not consider whether a person’s substance use or PLEs occurred first. Thus, we can only speculate about the causal role of substance use, including cannabis, in this relationship; alternative explanations for our findings include a shared genetic predisposition for substance use and PLEs, or the use of substances following PLEs to attenuate unwanted PLEs or other psychotic disorder-like symptoms (the “self-medication” hypothesis). Another limitation was the lack of available data on the earliest age of substance use and PLEs, as well as how many total PLEs a participant experienced. We also did not have access to data on potency of cannabis used; there is evidence that cannabis with a greater concentration of THC is more likely to result in psychotic symptoms and risk for psychotic disorders^4,6^, but we were unable to test this in our current study.

Our findings suggest that cannabis use and genetic risk for schizophrenia are correlated but separable risk factors for PLEs. Whether the association between cannabis use and PLEs is driven by cannabis use being a causal risk factor for PLEs or some other pathway (self-medication, shared genetic predisposition) or a combination of these mechanisms, our findings call for prospective studies on the potential impact of more frequent cannabis use on the development of PLEs. Our findings also support serious consideration of other substances as potential risk factors for PLEs in addition to cannabis. Given continued increases in the legalization of cannabis and the potency of cannabis sold, large longitudinal studies designed to thoroughly characterize the extent to which frequent use of cannabis and other substances leads to PLEs and potentially more serious psychopathology are sorely needed.

## Data Availability

All data analyzed in the present study are available to registered researchers via the All of Us Researcher Workbench.

## ACKNOWLEDGEMENTS

We gratefully acknowledge *All of Us* participants for their contributions, without whom this research would not have been possible. We also thank the National Institutes of Health’s *All of Us* <u>Research Program</u> for making available the participant samples examined in this study.

